# Quantifying the Risk of General Health and Early COVID-19 Spread in Residential Buildings with Deep Learning and Expert-augmented Machine Learning

**DOI:** 10.1101/2023.04.25.23289115

**Authors:** Jingjing Guan, Eman Leung, Kin On Kwok, Chi Tim Hung, Albert Lee, Ka Chun Chong, Carrie Ho Kwan Yam, Clement KM. Cheung, Hendrik Tieben, Hector W.H. Tsang, Eng-kiong Yeoh

## Abstract

Buildings’ built environment has been linked to their occupants’ health. It remains unclear whether those elements that predisposed its residents to poor general health before the two SARS pandemics also put residents at risk of contracting COVID-19 during early outbreaks. Relevant research to uncover the associations is essential, but there lacks a systematic examination of the relative contributions of different elements in one’s built environment and other non-environmental factors, singly or jointly. Hence, the current study developed a deep-learning approach with multiple input channels to capture the hierarchical relationships among an individual’s socioecology’s demographical, medical, behavioral, psychosocial, and built-environment levels. Our findings supported that 1) deep-learning models whose inputs were structured according to the hierarchy of one’s socioecology outperformed plain models with one-layered input in predicting one’s general health outcomes, with the model whose hierarchically structured input layers included one’s built environment performed best; 2) built-environment features were more important to general health compared to features of one’s sociodemographic and their health-related quality of life, behaviors, and service utilization; 3) a composite score representing built-environment features’ statistical importance to general health significantly predicted building-level COVID-19 case counts; and 4) building configurations derived from the expert-augmented learning of granular built-environment features that were of high importance to the general health were also linked to building-level COVID-19 case counts of external samples. Specific built environments put residents at risk for poor general health and COVID-19 infections. Our machine-learning approach can benefit future quantitative research on sick buildings, health surveillance, and housing design.

**Highlights:**

- The current modeling approaches for COVID-19 transmission at early spread are limited due to uncertainty and rare events.
- Socio-ecological structure (SES) can organize variables from different hierarchies of a total environment.
- TensorFlow-based deep learning with recurrent and convolutional neural networks is developed to explain general health with SES-organized variables.
- Among SES factors, built environments have a greater association with general health.
- Built-environment risks on individual general health associated with early-spread COVID-19 infections in residential buildings.

## 1. Introduction

More research efforts had been advocated for sick/healthy building research of residential, educational, and hospital buildings, instead of office buildings, with quantitative methods rather than subjective assessment through surveys [1]. The pandemics and other extreme events have been driving the effective collaboration between building practitioners, health professionals, and data scientists to develop a common vocabulary (definitions, metrics), data collection protocols, and analysis methods related to occupant health in buildings [1,2]. Machine learning methods were used to build risk prediction models for the sick building syndrome [3], but neither residential buildings nor diverse machine learning methods were widely studied yet. To fill the research gap, this study aimed to provide a quantitative approach to evaluating the role of known environmental and sociodemographic factors on residents’ health with deep learning approaches.

Elements from the internal built environment of residential buildings can put individuals at risk of respiratory diseases, tuberculosis, and influenza [4–6]. These elements are particularly relevant in very densely populated cities [7,8]. It has been shown that the built environment relates to not only residents’ physical health but also mental health, especially within communities with poor housing conditions [9]. Never did the impact of one’s internal built environment on health draw more attention from professionals and the public than during the Severe Acute Respiratory Syndrome outbreak in 2003 (hereafter, ‘SARS’), when the internal built environments of a residential estate (Amoy Gardens) and a hotel (Metropole Hotel) were responsible for the two community outbreaks in Hong Kong (HK) [10]. In both cases, the buildings’ configurations exacerbated the effects of the built environment on disease transmission. For example, the evidence was found that the “re-entrant bays” of buildings in Amoy Gardens facilitated wind-induced natural ventilation [11], allowing aerosol to enter from the sewage of SARS-infected residents [12]. On the other hand, the spatial connectivity of building configuration was associated with increased microbe diversity within a given space [13], which was reflected in the SARS transmission patterns in the Metropole Hotel’s layout [10].

Unlike SARS, COVID-19 (referring to coronavirus disease caused by severe acute respiratory syndrome coronavirus-2 (SARS-CoV-2) RNA hereafter) clusters were found in even more residential buildings in HK [14,15]. The environmental transmission was observed in COVID-19 cases [16]. With the solid evidence supporting the airborne transmission of COVID-19 in indoor air environments [17], a few studies have examined how various environmental systems in residential buildings served as the mechanisms for COVID-19 transmission. In particular, it was found that different ventilation systems had varying effects on COVID-19 transmission [18]. Most importantly, various elements in one’s built environment do not affect the transmission individually but do so as part of a building’s configuration, as previous literature on SARS transmission suggests. For instance, although the plumbing system and indoor airflow work together to spread aerosol from the sewage [19], transmission is also affected by how adjacent floors [19] and building blocks [20] are configured. As such, research into the mechanism of COVID-19 transmission has been extended beyond individual built elements’ contributions to examine the effect of their configuration in buildings. Another research stream has focused on building configuration that is conducive to spatial connectivity [13] and thus affects the effectiveness of social distancing measures and exacerbates ventilation’s effects on the person-to-person spread of COVID-19 [18,21,22]. Moreover, poor access to light has also been attributed to the SARS outbreak [23]. It is noteworthy that the same building configurations that were responsible for communicable diseases have also been linked to a family of symptoms associated with one’s general health with unspecific causes [24–27] and chronic illnesses [6,28,29]. There seem to be complex associations between various parameters of the built environment and residents’ health.

Specifically, to the best of our knowledge, no published study has directly tested the hypothesis that the same building elements or configurations are concurrently associated with poor general health and susceptibility to infectious diseases like COVID-19 among residents. Nor has any systematic investigation been conducted to isolate the effects of specific elements in one’s built environment on health outcomes from other potential sources of influence, such as one’s clinical status, social relationships, social institutions to which one is affiliated, and the policies that shape society. We noticed that the lack of a theoretical framework for formulating a model can be addressed by the United States’ Centers for Disease Control and Prevention (CDC)’s social-ecological framework, which has always held as its central tenet that different sources of influence in one’s ecosystem must all be addressed to prevent harm and poor health [30,31]. The CDC’s socio-ecological framework can be adapted to elucidate or prevent COVID-19’s initial outbreak and re-emergence since research suggests that one’s living environment could play a role within other influences in one’s ecosystem.

Although the CDC’s socio-ecological framework has informed the concepts regarding prevention targets and strategies, we have yet to find an empirical study that follows the framework’s multifaceted nature in identifying prevention targets across one’s socioecology and modeling their intricate influences on health outcomes accordingly. The lack of socio-ecological-framework-inspired empirical research may be attributable to the traditional statistical methods’ inability to model the hierarchical and intricate factors in one’s socioecology when there are insufficient sample sizes of unique units at each hierarchy. To fill this research gap, namely, to align our analytics with CDC’s layered and hierarchically organized socio-ecological framework, we developed a deep neural network model to analyze hierarchically-related parameters using multiple input channels, wherein each channel has its own processing unit (i.e., the multiple convolutional heads that constituted a Multi-Headed Hierarchical Convolutional Neural Network (MHHCNN; [32]) based on inputs processed by Long Short-Term Memory (LSTM) units [33]. Unsupervised-learning neural networks were used for feature extraction to recognize geographical patterns from satellite images to discern the spatial risk of COVID-19 and further explained it with the social determinant of health [34], which treated factors of the total environment as a plain structure. To sum up, the multilayered and diverse nature of socio-ecological contributors to COVID-19 was never examined, nor has the difference in COVID-19 risk among buildings of various architectural configurations co-locate in the same geographical area been studied. In addition, enhancing structural alignment with the sociological framework compared to traditional neural networks, MHHCNN also showed benefits beyond traditional neural networks’ ability to handle high-dimensional data with high volumes, sparsity, complexity, and potential endogeneity [35–37]. The additional benefits of MHHCNN are best illustrated in Canizo, Triguero, Conde, and Onieva (2019) [38]: 1) feature extraction is enhanced by focusing only on one particular input channel at a time rather than on all features at once; 2) each convolutional head can be adjusted to the specific nature of the data of each socioecological level; and 3) it makes the model architecture flexible to adapt it to new configurations with added socio-ecological levels as the convolutional heads can easily be added, modified or removed. Furthermore, an MHHCNN has better performance and a shorter convergence time than a traditional neural network [32].

However, notwithstanding their ability to handle complex and high-dimensional data and relationships, traditional neural networks, and deep learning algorithms, in general, often produce results that are: hard to interpret, overfitted and lacking generalizability, and are susceptible to clinical data’s inherent biases and confounds [39]. Recently, it has been noted that by augmenting the machine learning process with expert input (so-called expert-augmented learning or continual learning), the findings will be more interpretable and generalizable with fewer confounds and biases [39,40]. For the current study, we adapted the learning scenario that, after the neural network is trained, its parameters are optimized for the new task and no longer for the previous one [41]. The objective of the current study is to quantify various features’ risks of poor general health using MHHCNN and using expert-augmented learning to subtract the risk scores related to the built environment and evaluate its association with contracting COVID-19 among residents of 35 buildings in eight Hong Kong public housing estates during the early spread of SARS-CoV-2.

## 2. Hypotheses

Firstly, we hypothesized that the performance of modeling public housing residents’ general health outcomes from features representing the demographical, medical, behavioral, psychosocial, household characteristics and built environment of the participants’ socioecology would be improved by organizing input features in a hierarchical architecture using MHHCNN to align with the participants’ socioecology. Secondly, we hypothesized that public housing’s built environment features were of higher marginal statistical importance (as measured by Euclidian and cosine distances) to its residents’ general health than features representing residents’ demographics, psychosocial statuses, healthy behaviors, and medical history and health-related quality of life, and household characteristics. Thirdly, we hypothesized that the same set of built environment features contributed to building residents’ general health outcomes (parameterized as the feature importance assigned) and was also linked to the COVID-19 case counts accumulated in the same buildings from District A during the first 365 days of the pandemic. Finally, we also hypothesized that, through expert-augmented machine learning, the building configurations derived from granular built environment features of high importance to general health and COVID-19 in District A were also associated with COVID-19 case counts accumulated in District B during the same period.

## 3. Material and methods

### 3.1 Data

Data were integrated from multiple sources to capture different aspects of one’s socioecology. To examine the hypothesis that the same residential building elements that contributed to the residents’ poor health before COVID-19 also contributed to COVID-19 case counts, we analyzed data collected from a 2003 government-funded health assessment of District A’s residents and extracted the built environment features embedded in residents’ addresses in 2003 from HK Housing Authority’s database of Districts A and B during the same period when COVID-19 case counts of buildings from the studied districts’ public housing estates during the first wave of the pandemic in HK were downloaded from the Latest local situation of COVID-19 made public by the HK government and press releases.

Funded by the municipal government of District A and led by the regional public hospital network, a local NGO, and a university, a health assessment program was conducted between October 2003 and March 2004. The details of the sampling procedure for the health assessment and description of the sample were available elsewhere [42]. In short, one adult (18+) member of each household was randomly sampled from District A’s public housing estates’ buildings. One thousand seven hundred ninety-eight households were invited to participate in the study, resulting in 685 participants, 59% of whom were female. In addition, 25% of the sample is 65+, 15% are 50–59, 24% are 40–49, and 36% are aged 18–39 years old. Study participants were invited to provide their residential addresses (from which participant-level and building-level built environment features were matched by us) and data on their demographics, psychosocial statuses, healthy behaviors, medical history and medical quality of life, and household characteristics. The general health as the response variable for MHHCNN was a dichotomous measure and scored 1, if a study participant reported poor health, e.g., illness or feeling unwell, at the time of assessment, otherwise scored 0. The COVID-19 case counts in expert-augmented learning were measured at the building level. Built environment features were consistently measured among District A’s buildings where study participants lived and District B’s buildings that belonged to the most prominent public housing estate of the greatest amount of buildings and the longest history in a district adjacent to District A. Please refer to Table 1 for the list of outcome variables and predictive features ascertained from health assessments and other data sources.

**Table 1.**
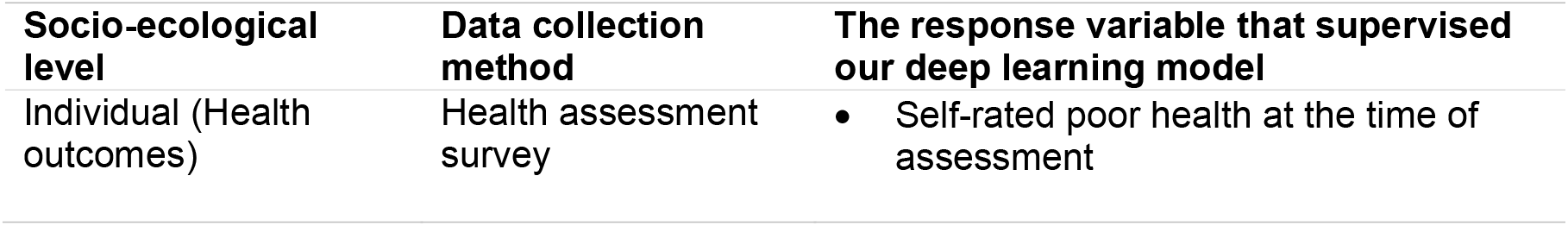
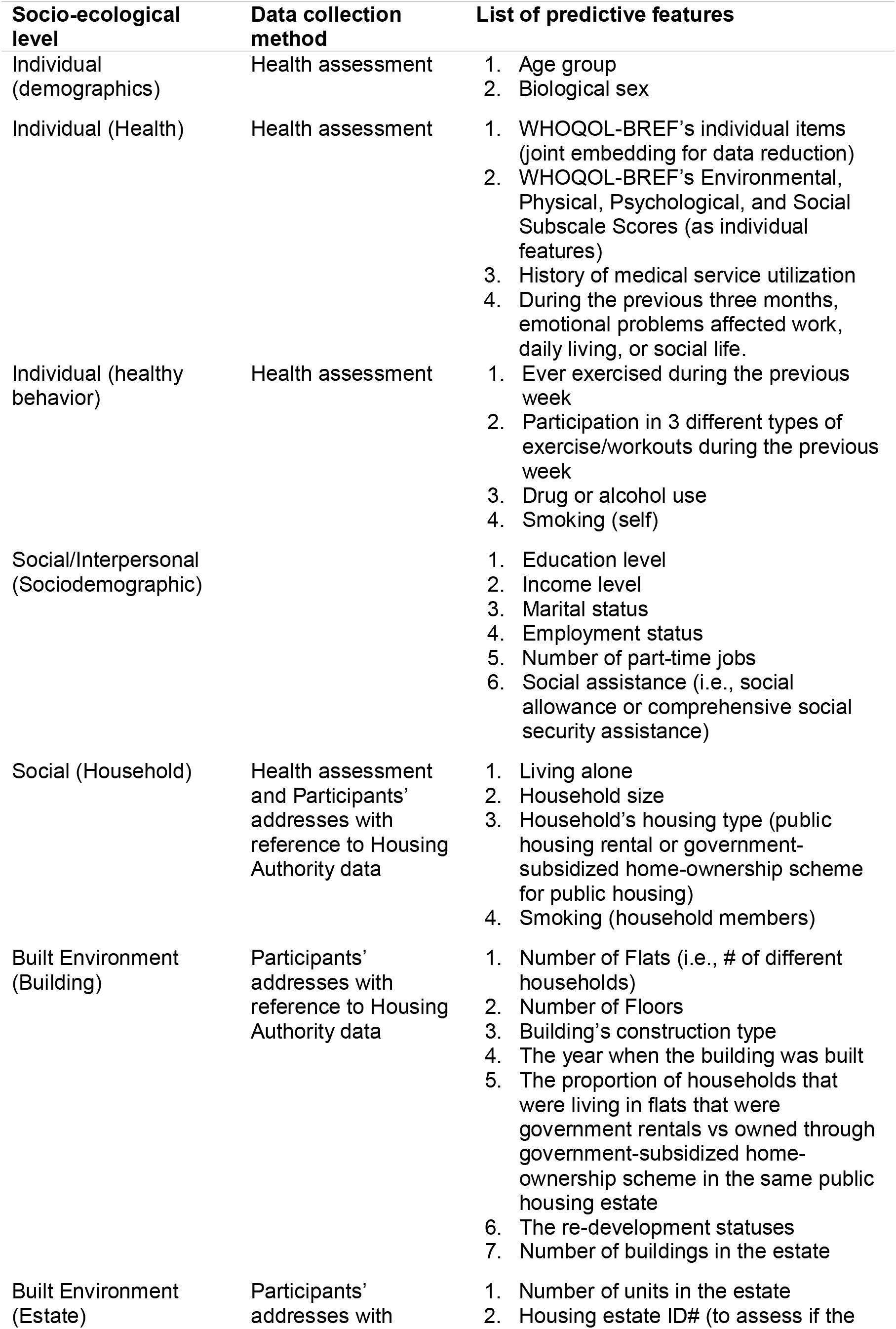
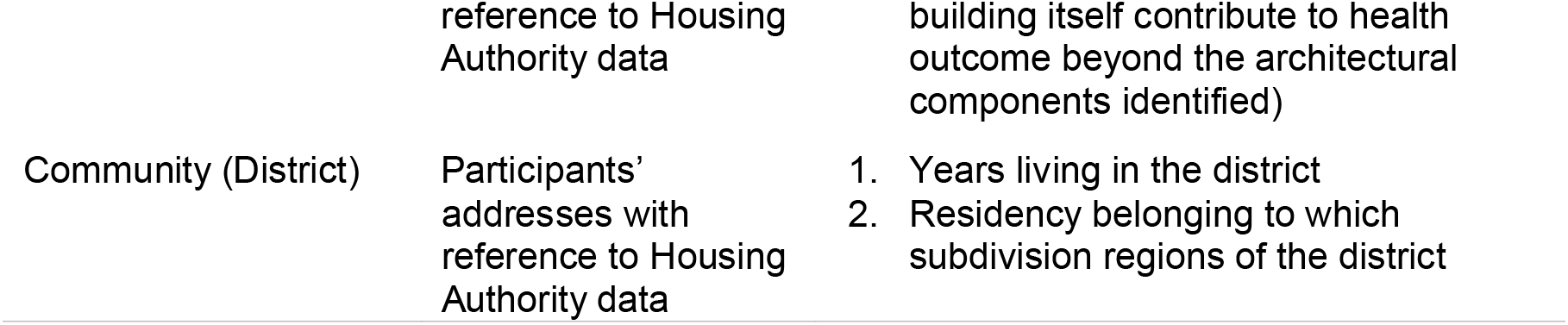
Data collected and their collection methods

### 3.2 Hypothesis testing

To examine if built environment features (in their granular forms or as modular configuration) that were of high statistical importance to residents’ general health outcomes according to MHHCNN were also linked to COVID-19 case counts accumulated in buildings whose residents reported their general health outcomes or in buildings located in an entirely different district, we tested our four hypotheses proposed above in the following steps.

We first examined the hypothesis that hierarchically structuring inputs representing one’s built environment and other aspects of one’s socioecology in MHHCNN’s architecture yields the most excellent performance in modeling general health outcomes. To this end, MHHCNN with a complete set of input features, including built environment ones, were compared to two single input-layered neural networks, with and without elements in one’s built environment as predictive features, respectively. In addition, the built environment-featured MHHCNN was also compared with an MHHCNN whose input lacked any built environment features. The architecture of MHHCNN is uniquely befitting for organizing separate streams of inputs that are hierarchically related. Each of MHHCNN’s input layers (i.e., each of the multiple processing ‘heads’ of CNNs that were hierarchically organized) was connected to an embedding layer followed by an LSTM layer (See Figure 1 for a schematic of its architecture; for the benefit of this hybrid convolutional and recurrent neural network approach, please see [43]). While each input layer constituted all levels of a single feature, different layers representing features belonging to the same socio-ecological hierarchy were concatenated and served as input to one convolutional layer. To assess and compare the performance of all models, we report below the area under the receiver operating curve (AUC) [44] and the number of epochs required for the models to converge.

**Figure 1.**
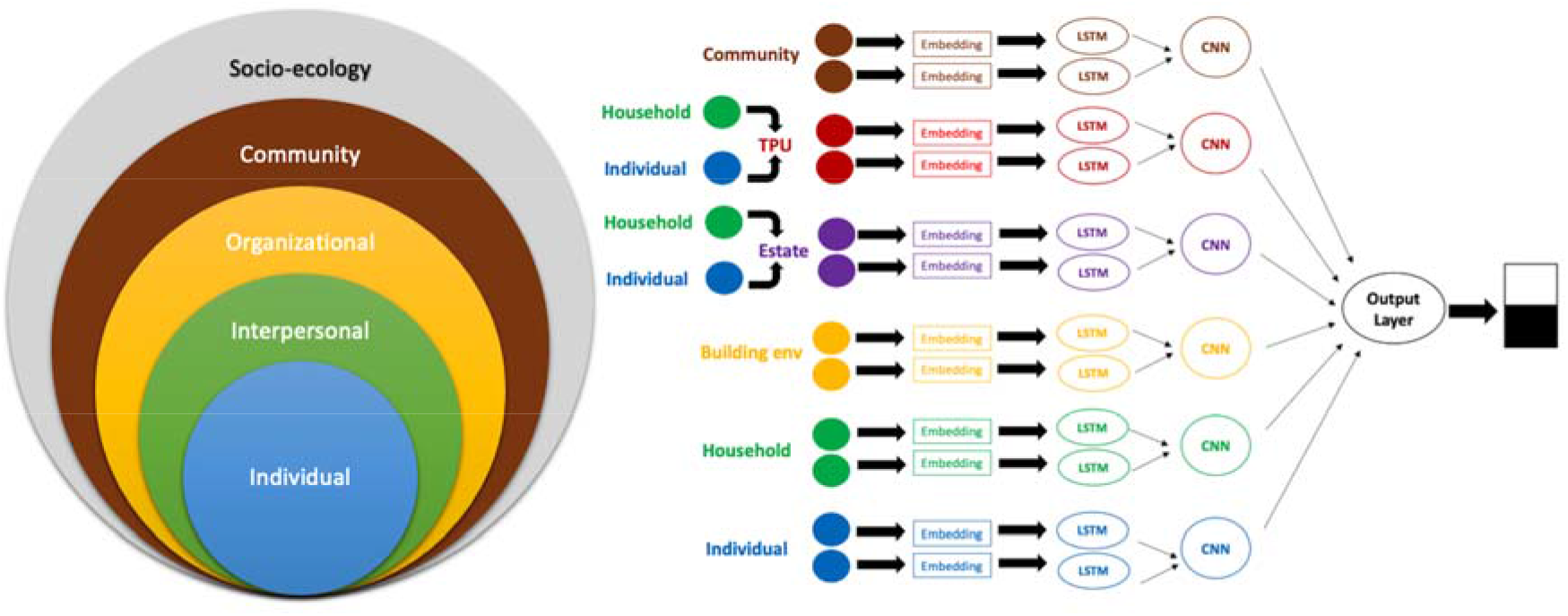
A schematic of our deep learning algorithm with multi-layered input from socio-ecological structured variables of the total environment of a community. (TPU: Tertiary Planning Units (TPUs) in the Population Census of HK. LSTM: Long Short-Term Memory layer. CNN: Convolutional Neural Network layer. Embedding: Embedding Input layer.)

Secondly, to quantify the unique contribution of one’s built environment to general health and to compare it with the contribution of other sociodemographic and health-related features, whose associations with general health were studied more frequently, MHHCNN was deployed to assign to each feature a value of statistical importance according to the magnitude of its unique contribution to general health outcomes (please refer to the appendix for how statistical importance for predictive features was estimated in the MHHCNN deployed in the current study). The unique contribution of each feature was calculated concerning the contributions of all other features and all possible interactions among neural network layers in our MHHCNN model.

Thirdly, to examine the hypothesis that the same built environment features contributing to residents’ general health are also responsible for the COVID-19 case count accumulated in each of the same buildings, a composite score was derived for each building by aggregating the building’s own value of statistical importance for each internal built environment feature identified by our model. For example, different buildings may differ in the number of flats on each floor and the total number of floors. Similarly, different buildings may have been built in different years, under different construction types, and for redevelopment purposes in some cases. Hence, in addition to estimating feature-level statistical importance, our model also assigns buildings that are different from each other for the same built environment feature different values of statistical importance for that built environment feature. Consequently, a building-level composite score was derived from aggregating the statistical importance of each granular built environment feature studied in each of the 35 buildings in District A to represent each studied building’s own built environment’s impact on its residents’ general health outcomes. Then, a Poisson regression was performed to predict COVID-19 case counts accumulated in each of the 35 buildings by the building-level composite score representing the same 35 buildings’ built environment. Below, the resulting incidence rate ratio was presented with its corresponding confidence interval and the p-value.

Finally, we tested the hypothesis that the built environment responsible for residents’ general health and COVID-19 in district A was also linked to COVID-19 case counts accumulated in District B’s buildings by first augmenting the granular built environment features of District A’s buildings into more generalizable modular features representing different configurations of buildings, before mapping the modular configurational features to the studied buildings in District B to predict the COVID-19 case counts they have accumulated during the first 365 days of the pandemic (see [45] for the benefit of feature modularization and other techniques of feature augmentation). Specifically, a panel of 6 academics from the School of Architecture of the lead authors’ institution was invited to review the granular built environment features’ contributions to general health outcomes and deduce the underlying building configurations responsible for the studied outcomes. Consequently, expert-augmented features representing different building configurations of high importance to general health outcomes were engineered. In addition, the effects of expert-augmented features on the general health outcomes of District A’s survey participants were tested again in an MHHCNN, which assigned statistical importance to each expert-augmented feature. Furthermore, the statistical importance given to each expert-augmented feature was then applied to calculate a composite score for each studied building in District B, whose architectural designs included different variants of all the building configurations identified by the experts. The resulting building-level composite scores on modular building configuration assigned to the studied buildings in District B then served as the independent variable for modeling District B’s buildings’ cumulative COVID-19 case counts using Poisson regression.

## 4. Results

We found that the proposed predicting method, MHHCNN with built environment features, best fits the general health outcome of individuals in district A. We also found that, on the building level, general health-related built environment risks were linked to case counts accumulated in COVID-19’s early spread.

In detail, the general health outcomes-supervised MHHCNN model with built environment features included as one of its input hierarchies yielded better performance than traditional neural networks and MHHCNN lacking built environment features. As Figure 2 shows, MHHCNN with a complete set of input features, including ones representing the built environment (blue line), converged at 95% AUC with much fewer epochs (10) compared to MHHCNN with no built environment features as input (green line). While neural networks (NNs) with single-layered input performed much worse by comparison, the single-layered NN with built environment features eventually reached 85% AUC at around the 90^th^ epoch.

**Figure 2.**
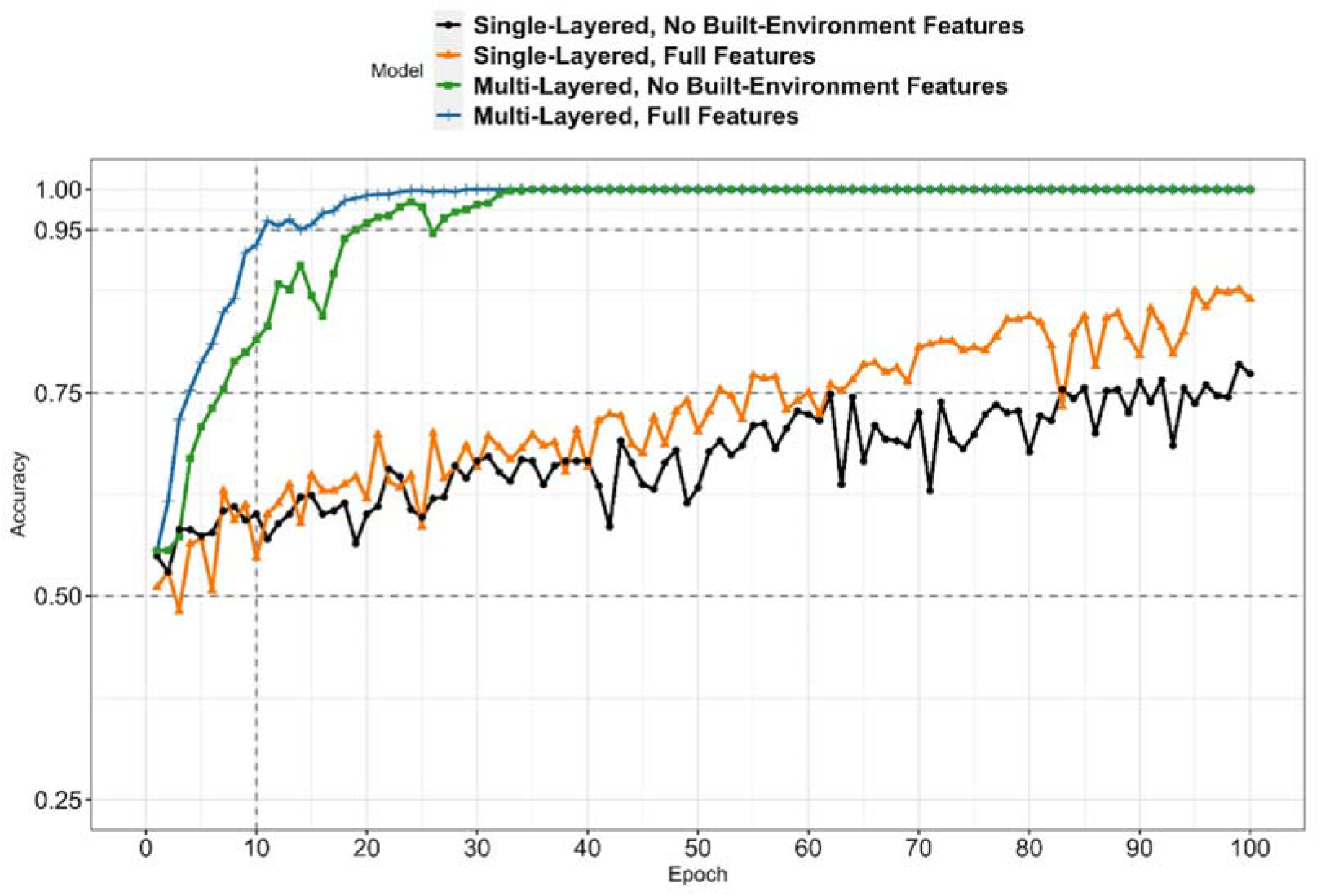
Validation AUCs of Single (flat) vs. Multiple Input Layers, with and without Built Environment Features.

Secondly, features representing public housing’s built environment were of higher marginal statistical importance to its residents’ general health than the majority of the features representing residents’ demographics, psychosocial statuses, healthy behaviors, and medical history. Figure 3 shows the marginal feature importance of the top 30 features, including features associated with the built environment, extracted from the MHHCNN. The higher the value of the feature’s marginal importance, the greater its unique contribution to one’s general health outcome with the contributions of all other predictive features and all the possible interactions among them. As the figure shows, on the list of features ordered in descending marginal statistical importance, external built environment features, such as the subdivision of the regions in which the building was located and the number of buildings in an estate, ranked 2^nd^ and 5^th^, respectively. The Internal built environment features, such as the average number of flats (representing the number of unique households) per floor and the number of floors in a building, ranked 4^th^ and 7^th^, respectively. In addition, among the top 30 features with the greatest marginal statistical importance, the year the buildings were built, the construction type of the buildings, and the re-development status of the buildings were ranked 23^rd^, 25^th^, and 30^th^, respectively.

**Figure 3.**
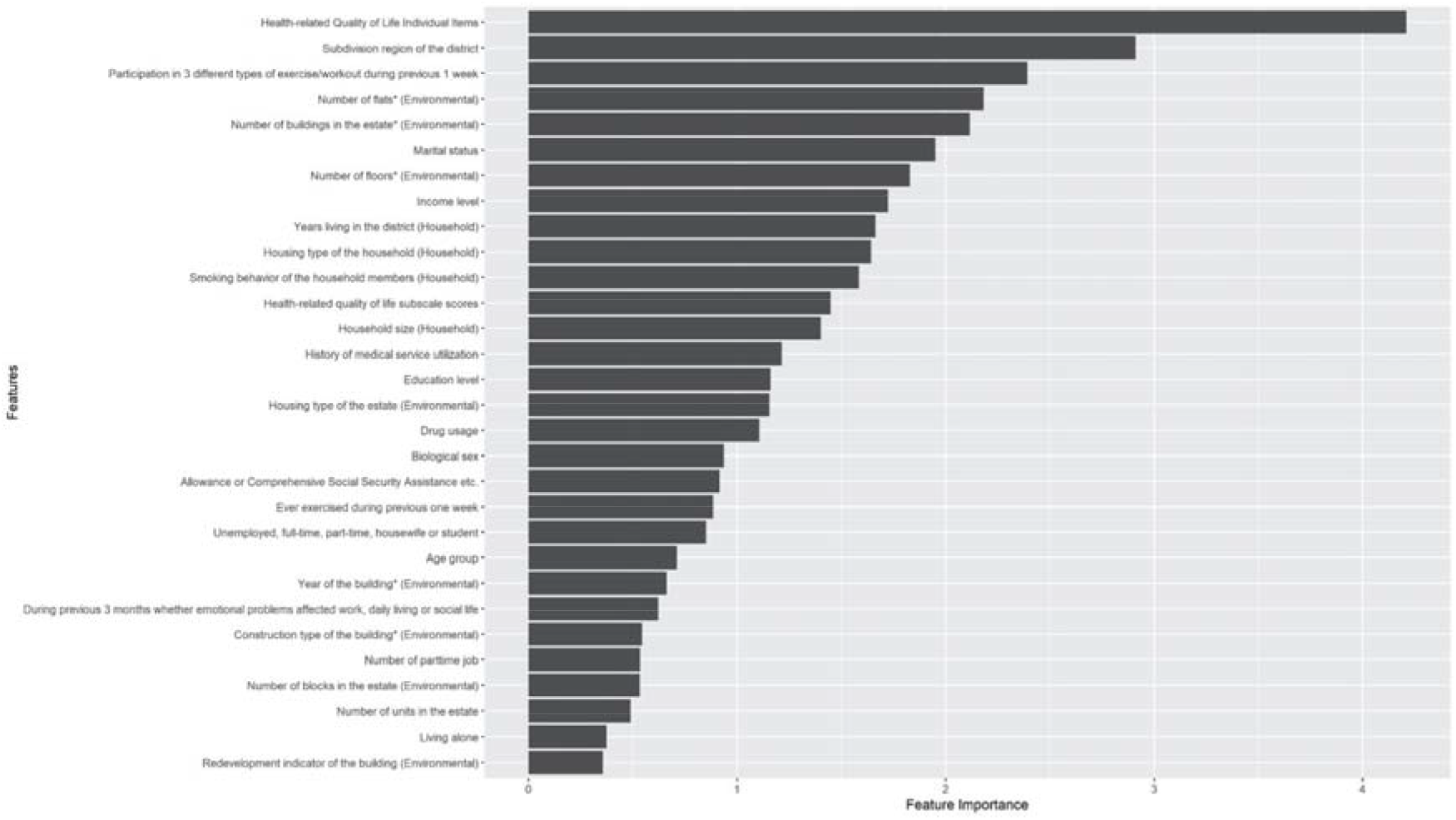
Marginal Importance of Predictive Features for MHHCNN.

Thirdly, built environment features that contributed to general health outcomes were also linked to COVID-19 case counts accumulated in the same buildings from District A during the first wave of the pandemic. Table 2 shows the statistical importance of each granular internal built environment feature studied in each of the 35 buildings in District A, the corresponding building-level composite score, and the cumulative COVID-19 case counts. Poisson regression revealed that the composite score that represented the aggregated internal built-environment features’ importance to general health outcomes was significantly associated with first-365-day cumulative COVID-19 case counts of the same buildings with an incidence rate ratio of 5.83 (95% CI: 3.03-10.91; p-value <.001).

**Table 2.** Built Environment Elements’ Feature Importance, Composite Score, and the Number of Confirmed COVID-19 Case Counts of Each Study Building in District A.

| Building ID | # of Flats | # of Floors | Year of Building | Construction Type | Re-development | Composite score of the five elements | # of COVID-19 Confirmed Cases |
| --- | --- | --- | --- | --- | --- | --- | --- |

|  |  |  |  |  |  |  | (Residential) |
| --- | --- | --- | --- | --- | --- | --- | --- |
| A | 1.318 | 0.180 | 0.000 | 0.000 | 0.016 | 1.514 | 13 |
| B | 0.597 | 0.378 | 0.007 | 0.000 | 0.003 | 0.985 | 6 |
| C | 0.448 | 0.393 | 0.000 | 0.000 | 0.002 | 0.843 | 5 |
| D | 0.660 | 0.182 | 0.000 | 0.000 | 0.016 | 0.858 | 4 |
| E | 0.287 | 0.490 | 0.000 | 0.012 | 0.002 | 0.791 | 3 |
| F | 0.698 | 0.534 | 0.000 | 0.022 | 0.000 | 1.254 | 3 |
| G | 0.922 | 0.738 | 0.000 | 0.022 | 0.000 | 1.682 | 2 |
| H | 0.178 | 0.381 | 0.000 | 0.000 | 0.000 | 0.559 | 2 |
| I | 0.740 | 0.155 | 0.000 | 0.000 | 0.002 | 0.897 | 2 |
| J | 0.557 | 0.276 | 0.007 | 0.015 | 0.003 | 0.858 | 2 |
| K | 0.064 | 0.180 | 0.000 | 0.000 | 0.000 | 0.244 | 2 |
| L | 0.234 | 0.195 | 0.064 | 0.015 | 0.000 | 0.508 | 2 |
| M | 0.314 | 0.222 | 0.000 | 0.000 | 0.000 | 0.536 | 2 |
| N | 0.742 | 0.126 | 0.064 | 0.000 | 0.000 | 0.932 | 1 |
| O | 0.293 | 0.287 | 0.000 | 0.000 | 0.002 | 0.582 | 1 |
| P | 0.514 | 0.404 | 0.023 | 0.000 | 0.002 | 0.943 | 1 |
| Q | 0.422 | 0.200 | 0.000 | 0.000 | 0.016 | 0.638 | 1 |
| R | 0.260 | 0.333 | 0.000 | 0.000 | 0.000 | 0.593 | 1 |
| S | 0.337 | 0.260 | 0.087 | 0.000 | 0.000 | 0.684 | 1 |
| T | 0.408 | 0.204 | 0.000 | 0.000 | 0.000 | 0.612 | 1 |
| U | 0.252 | 0.125 | 0.000 | 0.000 | 0.000 | 0.377 | 1 |
| V | 0.314 | 0.138 | 0.000 | 0.000 | 0.002 | 0.454 | 1 |
| W | 0.191 | 0.210 | 0.000 | 0.000 | 0.000 | 0.401 | 1 |
| X | 0.952 | 0.212 | 0.000 | 0.017 | 0.002 | 1.183 | 0 |
| Y | 0.577 | 0.205 | 0.000 | 0.000 | 0.002 | 0.784 | 0 |
| Z | 0.371 | 0.393 | 0.007 | 0.015 | 0.003 | 0.789 | 0 |
| AA | 0.439 | 0.360 | 0.000 | 0.000 | 0.000 | 0.799 | 0 |
| BB | 0.270 | 0.482 | 0.023 | 0.000 | 0.002 | 0.777 | 0 |
| CC | 0.333 | 0.333 | 0.000 | 0.017 | 0.002 | 0.685 | 0 |
| DD | 0.313 | 0.333 | 0.007 | 0.015 | 0.003 | 0.671 | 0 |
| EE | 0.405 | 0.254 | 0.000 | 0.000 | 0.000 | 0.659 | 0 |
| FF | 0.292 | 0.356 | 0.000 | 0.000 | 0.000 | 0.648 | 0 |
| GG | 0.310 | 0.204 | 0.000 | 0.000 | 0.018 | 0.532 | 0 |
| HH | 0.337 | 0.121 | 0.000 | 0.000 | 0.000 | 0.458 | 0 |
| II | 0.207 | 0.000 | 0.000 | 0.024 | 0.000 | 0.231 | 0 |

Finally, building configurations derived from the expert-augmented learning of granular built environment features of high importance to the general health of residents of District A’s buildings were linked to COVID-19 case count accumulated in the studied buildings of both District A and B. Table 3 shows the statistical importance of building configurations derived from the expert-augmented learning of granular built environment features of high importance to the general health of residents of District A’s buildings being mapped onto the studied buildings in District B. The three modular features of building configuration were 1) being connected to other buildings, 2) being part of a building block, and 3) construction types. Table 3 also shows the composite scores derived from aggregating the three modular features of building configuration and the cumulative COVID-19 case counts observed in the studied buildings in District B. Poisson regression revealed that the composite score representing the aggregation of the feature importance of the three modular features of building configuration identified from District A’s buildings studied predicted the cumulative COVID-19 case counts of the 18 buildings in District B’s most prominent public housing estate onto which the modular configurational features could be mapped (incidence rate ratio=450.02 (95% CI: 24.47-17913.29; p-value <.001)).

**Table 3.** Out-sample Building Configurations’ Feature Importance, Composite Score, and the Number of Confirmed COVID-19 Case Counts of Buildings from the Most Prominent Public Housing Estate in District B.

| Building ID | Connected to Other Building | Part of a Building Block | Construction Type | Composite Score | # of COVID-19 Confirmed Cases (Residential) |
| --- | --- | --- | --- | --- | --- |
| 1 | 0.187 | 0.295 | -0.004 | 0.478 | 7 |
| 2 | 0.013 | 0.295 | -0.004 | 0.304 | 5 |
| 3 | 0.013 | 0.295 | -0.004 | 0.304 | 3 |
| 4 | 0.187 | 0.295 | 0.017 | 0.499 | 2 |
| 5 | 0.187 | -0.076 | -0.015 | 0.096 | 1 |
| 6 | 0.013 | 0.295 | -0.004 | 0.304 | 0 |
| 7 | 0.187 | 0.295 | -0.004 | 0.478 | 0 |
| 8 | 0.013 | 0.295 | -0.004 | 0.304 | 0 |
| 9 | 0.013 | 0.295 | -0.004 | 0.304 | 0 |
| 10 | 0.013 | 0.295 | -0.004 | 0.304 | 0 |
| 11 | 0.013 | -0.076 | 0.017 | -0.046 | 0 |
| 12 | 0.013 | -0.076 | 0.017 | -0.046 | 0 |
| 13 | 0.013 | -0.076 | 0.017 | -0.046 | 0 |
| 14 | 0.013 | -0.076 | 0.017 | -0.046 | 0 |
| 15 | 0.013 | -0.076 | 0.017 | -0.046 | 0 |
| 16 | 0.013 | -0.076 | 0.017 | -0.046 | 0 |
| 17 | 0.013 | -0.076 | 0.017 | -0.046 | 0 |
| 18 | 0.013 | -0.076 | -0.023 | -0.086 | 0 |

## 5. Discussion

Methodological, statistical, and computational significance was found with the proposed approach in the four hypotheses we sought to test. First, socio-ecological structure and MHHCNN together served as a good match for organizing multi-hierarchies of variables into explaining general health outcomes, as our analysis revealed. The MHHCNN helps to address the methodological gap that a shortage of sample sizes but the large number of variables in each socio-ecological hierarchy limits the use of classic multilevel models in statistics, which is designated for handling multi-hierarchies structured data [46,47]. The proposed MHHCNN approach can handle data of multi-hierarchy nature that are commonly seen in the anthroposphere. The current study also echoes a recent study that developed a quantitative framework for evaluating floods/droughts management and the catchment from a socio-ecological system perspective [48]. Our MHHCNN goes one step further to implement the quantifiable framework with real-world data and tests its validity with new data that didn’t involve in the model training to increase confidence in the findings.

Specifically, by comparing the performance of built environment-featured MHHCNN with the version of MHHCNN that lacks built environment features and the implementation of single-input-layered neural networks, we can infer the superiority of the built environmentfeatured MHHCNN model. It is worth noting that, unlike the gap in performance between the two traditional neural networks or between the traditional neural networks and the MHHCNNs, the performance of the two MHHCNNs eventually converged. Nevertheless, the performance of the built environment-featured MHHCNN surpassed 95% AUC at around ten epochs earlier than the MHHCNN that lacked any built environment features.

In support of our second hypothesis tested, evidence showed that features representing public housing’s built environment were of higher marginal statistical importance to its residents’ general health than most features representing residents’ demographics, psychosocial statuses, healthy behaviors, and medical history, with healthrelated quality of life being a notable exception.

Our model had assigned great marginal importance to the built environment’s contribution to general health outcomes, even after accounting for the well-documented effects of residents’ sociodemographic and their health-related quality of life, behavior, and service utilization in the past. In addition to the constructional elements representing one’s internal built environment, such as the number of distinct households per floor and the number of flats in the buildings, other built environment features representing one’s living environment in a broader sense have also been assigned top-ten importance by our model. For example, the subdivision regions of the district where the residential building is located, the number of buildings in the estate, the number of years living in the neighborhood, and the household’s housing type (rental or owned via government subsidies). In addition, the effects of the built environment on general health outcomes cannot be attributed to population density alone, given that the model has already accounted for the contribution of the size of the household and the number of units and blocks in the estates, even though, unlike the similar population density measure of the number of buildings in the estate, these features had not been selected into the top ten.

Unlike the single-family detached or attached houses, which are typical residential constructions in some areas of the world, the current study focused on multistory apartment-based residential buildings, which are more high-rise, e.g., have thirteen floors or more. The MHHCNN and experts recognized the reported patterns but the current study hasn’t investigated the underlying causes of why such built environments had been more associated with adverse health outcomes. Hence, it is recommended that future research on one’s built environment should focus beyond these constructional elements and examine how they interact with other factors to contribute to the residents’ health outcomes. It is also recommended to investigate more buildings located in different subdivisions of the districts that differ in social-economic status and other elements in the external built environment, which may, in turn, contribute to the difference in health outcomes among residents of these buildings as a function of the years of living in the same environment.

Under the framework of the socio-ecological system, the findings reported here also support that specific built environment features that contributed to residents’ general health outcomes were also linked to COVID-19 case counts accumulated in the same buildings from District A during the first wave of the pandemic. The specific built environments found to be risky for residents’ general health and infection in the early spread of COVID-19 include being connected to other buildings directly, disregarding the connected buildings’ functions (e.g., residential or commercial); being part of a building block composed of highrise residential buildings usually; and specific construction types that are standardized in HK. Experts summarized the three granular internal built environment features: 1) being part of a building block, 2) being connected to other buildings, and 3) construction types. The variability in the building configurations our expert panel identified was associated with the differences in the studied buildings’ lighting, air ventilation, and connectivity – which previous research had linked to poor general health and SARS among the residents of HK public housing. Under the theoretical framework of the socio-ecological system, the current study newly quantitatively measured the effects of different building configurations on the number of reported infectious cases in the early stage of a pandemic. On the one hand, our findings can enlighten a way to study how to stop the spread of a pandemic outbreak at its early stage by tracing infectious cases according to specific built environment features. On the other hand, for long-term urban planning, more in-depth expert-augmented machine learning can be enabled with our approach to identifying potentially health-compromising building configurations in other public housing estates in different HK districts where there is yet any research linking the granular elements of one’s internal built environment to health outcome to inform tertiary-wide housing policies and regional health service planning and deployment.

Given that public housing’s internal built environment in HK is standardized across the tertiary with only limited variations, and given our findings that the effect of the built environment on health outcomes is persistent, the current study can therefore be relevant to long-term housing and healthcare policy in HK or places similarly densely populated as HK. Our findings echo the research on building unevenness of air exchange and pollution exchange. One public housing estate in HK is typically composed of multiple construction types and non-uniform building heights, which increased the variation of building unevenness. Recent research found that increased building unevenness complicated the canyon airflow structure and aggravated the pollution dispersion of compact urban blocks [49]. Another perspective is, historically speaking, in HK, most public housing’s architectural designs of the internal (constructional) built environment were dated before 1984. The stability of the internal built environment design could be the underlying reason why not only were selected elements of the internal built environment linked to the residents’ general health outcome assessed in the 2003 community diagnosis, but the internal built environment features’ importance to residents’ general health was also linked to the corresponding buildings’ COVID-19 case counts in 2020. In contrast to the stable internal (construction) built environment, the external built environment of public housing across HK has changed considerably. Hence, to ensure that the elements of the built environment whose effects on general health being studied were equivalent to the elements of the built environment to which such effects on general health were applied to predict COVID-19 case counts, we only aggregated the internal built environment features’ statistical importance to calculate the composite score. The evidence seems to point to the inference that the built environments of HK’s public housing have prolonged and evitable effects on its residents’ communicable and non-communicable disease diagnoses, not until the current study that we can properly attribute the effects quantitatively to corresponding environmental elements.

Finally, our findings also supported the hypothesis that building configurations derived from the expert-augmented learning of granular built environment features of high importance to the general health of residents of District A’s buildings were linked to COVID-19 case count accumulated in the studied buildings of both Districts A and B. Despite machine learning algorithms’ successes in modeling complex and intricate relationships across several disciplines, they often fail to generalize to new cases – especially when they are built from, or applied to, a small volume of data, given their data inefficiency [39]. Most importantly, it has been noted that, due to the lack of general knowledge in a specific domain, current machine learning algorithms often make oversights that appear trivial to experts but, at the same time, are of grave consequences if decisions were made based on such oversights, especially in healthcare [39]. Iteratively combining computational tools with human expertise holds promise for the identification of treatments for rare and neglected diseases, e.g., drugs for the COVID-19 [50] and optimizing hemodynamic predictors of spinal cord injury outcome [51], in areas of biological discovery where relevant data may be lacking or hidden in the mass of available biomedical literature [50]. Recently, expert-augmented machine learning has been advanced to gain the trust of artificial intelligence in the area of healthcare, e.g., [39] and environmental science, e.g., [52] by rectifying machine learning algorithms’ ungeneralizable findings, inefficient data usage, and the lack of domain-specific general knowledge. Expert-augmented machine learning in the current study sampled a smart and efficient dataset to study rare events (i.e., the building-level case counts during the early COVID-19 spread) with the aid of individual samples that shared the same total environment but never went through a pandemic, i.e., SARS or COVID-19. We tackled an alternate solution to study rare events, which could have been difficult to trace and the literature has been working on sampling or simulation techniques to study them, e.g., [53]. With the socio-ecological framework, deep learning model, and expert-augmented learning, individual-level information is of higher resolution than the building level which is like a stretching net to capture the useful information from step to step. In the research stream of COVID-19 studies, the proposed method helps to capture the uncertainty of COVID-19 transmission in the total built environment. In fact, owing to the dynamics of the nonlinear pandemic prediction model [54] and the small case counts, modeling and tracing COVID-19 transmission at the early stage of spread remains challenging. We not only established an approach from the data selection, the socio-ecological system as the theoretical framework, and an effective learning approach of both deep neural networks and expert-augmented learning, to quantify the relationship between internal built environment features’ importance to the general health of residents of the studied buildings in District A and these same buildings’ cumulative COVID-19 case counts but also generalized and validated the relationship to another sample of buildings in District B to predict the COVID-19 case counts these District B buildings had accumulated.

## 6. Limitations

In addition to the small number of buildings in Districts A and B studied, which the current study attempted to rectify with expert-augmented machine learning, another limitation of the current study was that the health needs assessment was only administered to public housing residents. Public health and housing policies have been changing since 2003. Nonetheless, the study identified significant relationships between building designs before the’90s, health assessments conducted in 2003 (before two SARS-related pandemics), and cumulative case counts of COVID-19 recorded in districts in 2020. The results suggest that the built environment’s effect on health outcomes may be persistent across time and generalizable across geographical locations in HK. The effects of the built environment on health outcomes reported here may not be generalizable to private housing residents as there is no consistent standard for private building designs in HK. Regarding social determinants of health, while private housing rental and ownership generally represent higher socio-economic status, the most vulnerable population in HK, even though a minority, are living in older private flats subdivided into several small units with a shared kitchen and bathroom. In addition to the well-documented poor physical and mental health of occupants in subdivided flats, the elevated risk of infectious diseases is also noteworthy. This study does not capture the adverse health impacts of such design layouts on the occupants. Furthermore, as private housing is not included in the study, we cannot accurately trace the potential spread of infectious diseases in a district; private and public housing are built close to one another in HK and share much of the public space, and some are even co-located.

By including only elements of one’s internal built environment in predicting general health and COVID-19 outcomes, we may have undermined the actual effect of one’s built environment. For example, among those internal built environment features whose importance to general health was aggregated into a composite score, only the number of flats and floors were of top-ten marginal importance to the studied general health outcomes. Nevertheless, the remaining internal built environment features whose statistical importance was aggregated into the composite score, such as the year a building was built, the construction type of the building, and whether the building had undergone re-development, ranked 23rd, 25th, and 30th, respectively, in terms of their marginal importance to the resident’s general health. On the other hand, features representing both the external and internal environment were of top-ten importance to the building residents’ general health outcome even after accounting for the contributions (singly or jointly) of other top-ten features such as health-related quality of life, exercise, material status, and income. Notably, the unique contribution of the number of flats at the estate level (i.e., the number of individual households living in the same estate) was greater than, and independent of the contribution of features related to population density. In other words, general health is affected by the number of individual families living in the buildings or estates, not only the size of each family or the total number of residents. This finding is consistent with research on the risk of communicable diseases in cross-family contact.

## Data Availability

All data produced in the present study are not available to be shared by the authors

## Abbreviations

AUC: area under the receiver operating curve.
CDC: The Centers for Disease Control and Prevention of the United States.
CNN: convolutional neural network.
COVID-19: Coronavirus disease 2019 caused by severe acute respiratory syndrome coronavirus-2.
LSTM: Long short-term memory.
MHHCNN: Multi-headed hierarchical convolutional neural network. NN: neural network.
SARS: Severe Acute Respiratory Syndrome. SES: Socio-ecological structure.
TPU: Tertiary Planning Unit.

## Acknowledgment

The work was supported by the Hong Kong government’s Strategic Public Policy Research Funding Scheme [grant number S2019.A4.015.19S].

The authors declare that they have no known competing financial interests or personal relationships that could have appeared to influence the work reported in this paper.

## Notes

### Competing Interest Statement

The authors have declared no competing interest.

### Funding Statement

This study was funded by the Hong Kong government Strategic Public Policy Research Funding Scheme [grant number S2019.A4.015.19S].

### Author Declarations

Ethics committee/IRB of the Chinese University of Hong Kong gave ethical approval for this work

## REFERENCES

[1] M. Awada, B. Becerik-Gerber, E. White, S. Hoque, Z. O’Neill, G. Pedrielli, J. Wen, T. Wu, Occupant health in buildings: Impact of the COVID-19 pandemic on the opinions of building professionals and implications on research, Build. Environ. 207 (2022) 108440. https://doi.org/https://doi.org/10.1016/j.buildenv.2021.108440.

[2] M. Awada, B. Becerik-Gerber, S. Hoque, Z. O’Neill, G. Pedrielli, J. Wen, T. Wu, Ten questions concerning occupant health in buildings during normal operations and extreme events including the COVID-19 pandemic, Build. Environ. 188 (2021) 107480. https://doi.org/https://doi.org/10.1016/j.buildenv.2020.107480.

[3] L. Fan, Y. Ding, Research on risk scorecard of sick building syndrome based on machine learning, Build. Environ. 211 (2022) 108710. https://doi.org/https://doi.org/10.1016/j.buildenv.2021.108710.

[4] Y. Li, G.M. Leung, J.W. Tang, X. Yang, C.Y.H. Chao, J.Z. Lin, J.W. Lu, P. V. Nielsen, J. Niu, H. Qian, A.C. Sleigh, H.J.J. Su, J. Sundell, T.W. Wong, P.L. Yuen, Role of ventilation in airborne transmission of infectious agents in the built environment – A multidisciplinary systematic review, Indoor Air. 17 (2007) 2–18. https://doi.org/10.1111/j.1600-0668.2006.00445.x.

[5] T. Wirth, URBAN NEGLECT: THE ENVIRONMENT, PUBLIC HEALTH, AND INFLUENZA IN PHILADELPHIA, 1915-1919, Pennsylvania Hist. A J. Mid-Atlantic Stud. 73 (2006) 316–342. http://www.jstor.org/stable/27778742.

[6] N. Pinter-Wollman, A. Jelic, N.M. Wells, The impact of the built environment on health behaviours and disease transmission in social systems, Philos. Trans. R. Soc. B Biol. Sci. 373 (2018). https://doi.org/10.1098/rstb.2017.0245.

[7] E. Ng, Policies and technical guidelines for urban planning of high-density cities – air ventilation assessment (AVA) of Hong Kong, Build. Environ. 44 (2009) 1478–1488. https://doi.org/10.1016/j.buildenv.2008.06.013.

[8] J. Kandt, S.-S. Chang, P. Yip, R. Burdett, The spatial pattern of premature mortality in Hong Kong: How does it relate to public housing?, Urban Stud. 54 (2016) 1211–1234. https://doi.org/10.1177/0042098015620341.

[9] Z. Kan, M.P. Kwan, M.K. Ng, H. Tieben, The Impacts of Housing Characteristics and Built-Environment Features on Mental Health, Int. J. Environ. Res. Public Health. 19 (2022). https://doi.org/10.3390/ijerph19095143.

[10] C.R. Braden, S.F. Dowell, D.B. Jernigan, J.M. Hughes, Progress in global surveillance and response capacity 10 years after severe acute respiratory syndrome., Emerg. Infect. Dis. 19 (2013) 864–869. https://doi.org/10.3201/eid1906.130192.

[11] C.K.C. Cheng, K.M. Lam, Y.T.A. Leung, K. Yang, H.W. Li Danny, C.P. Cheung Sherman, Wind-induced natural ventilation of re-entrant bays in a high-rise building, J. Wind Eng. Ind. Aerodyn. 99 (2011) 79–90. https://doi.org/10.1016/j.jweia.2010.11.002.

[12] I.T.S. Yu, Y. Li, T.W. Wong, W. Tam, A.T. Chan, J.H.W. Lee, D.Y.C. Leung, T. Ho, Evidence of airborne transmission of the severe acute respiratory syndrome virus., N. Engl. J. Med. 350 (2004) 1731–1739. https://doi.org/10.1056/NEJMoa032867.

[13] S.W. Kembel, J.F. Meadow, T.K. O’Connor, G. Mhuireach, D. Northcutt, J. Kline, M. Moriyama, G.Z. Brown, B.J.M. Bohannan, J.L. Green, Architectural Design Drives the Biogeography of Indoor Bacterial Communities, PLoS One. 9 (2014) e87093. https://doi.org/10.1371/journal.pone.0087093.

[14] C.K.C. Lai, R.W.Y. Ng, M.C.S. Wong, K.C. Chong, Y.K. Yeoh, Z. Chen, P.K.S. Chan, Epidemiological characteristics of the first 100 cases of coronavirus disease 2019 (COVID-19) in Hong Kong Special Administrative Region, China, a city with a stringent containment policy, Int. J. Epidemiol. 49 (2020) 1096–1105. https://doi.org/10.1093/ije/dyaa106.

[15] K.Y. Lai, C. Webster, S. Kumari, C. Sarkar, The nature of cities and the Covid-19 pandemic., Curr. Opin. Environ. Sustain. 46 (2020) 27–31. https://doi.org/10.1016/j.cosust.2020.08.008.

[16] S. Vardoulakis, M. Sheel, A. Lal, D. Gray, COVID-19 environmental transmission and preventive public health measures, Aust. N. Z. J. Public Health. 44 (2020) 333–335. https://doi.org/10.1111/1753-6405.13033.

[17] Z. Noorimotlagh, N. Jaafarzadeh, S.S. Martínez, S.A. Mirzaee, A systematic review of possible airborne transmission of the COVID-19 virus (SARS-CoV-2) in the indoor air environment., Environ. Res. 193 (2021) 110612. https://doi.org/10.1016/j.envres.2020.110612.

[18] R.K. Bhagat, M.S.D. Wykes, S.B. Dalziel, P.F. Linden, Effects of ventilation on the indoor spread of COVID-19, J. Fluid Mech. 903 (2020) F1. https://doi.org/10.1017/jfm.2020.720.

[19] M. Gormley, T.J. Aspray, D.A. Kelly, C. Rodriguez-Gil, Pathogen cross-transmission via building sanitary plumbing systems in a full scale pilot test-rig., PLoS One. 12 (2017) e0171556. https://doi.org/10.1371/journal.pone.0171556.

[20] C. Yip, W.L. Chang, K.H. Yeung, I.T.S. Yu, Possible meteorological influence on the Severe Acute Respiratory Syndrome (SARS) community outbreak at Amoy Gardens, Hong Kong, J. Environ. Health. 70 (2007) 39–46. https://www.jstor.org/stable/26327426.

[21] C. Sun, Z. Zhai, The efficacy of social distance and ventilation effectiveness in preventing COVID-19 transmission, Sustain. Cities Soc. 62 (2020) 102390. https://doi.org/10.1016/j.scs.2020.102390.

[22] H. Dai, B. Zhao, Association of the infection probability of COVID-19 with ventilation rates in confined spaces., Build. Simul. 13 (2020) 1321–1327. https://doi.org/10.1007/s12273-020-0703-5.

[23] Y. Li, S. Duan, I.T.S. Yu, T.W. Wong, Multi-zone modeling of probable SARS virus transmission by airflow between flats in Block E, Amoy Gardens, Indoor Air. 15 (2005) 96–111. https://doi.org/10.1111/j.1600-0668.2004.00318.x.

[24] A.P. Jones, Indoor air quality and health, Atmos. Environ. 33 (1999) 4535–4564. https://doi.org/10.1016/S1474-8177(02)80006-7.

[25] P. Carrer, P. Wargocki, A. Fanetti, W. Bischof, E. De Oliveira Fernandes, T. Hartmann, S. Kephalopoulos, S. Palkonen, O. Seppänen, What does the scientific literature tell us about the ventilation–health relationship in public and residential buildings?, Build. Environ. 94 (2015) 273–286. https://doi.org/https://doi.org/10.1016/j.buildenv.2015.08.011.

[26] S. Deitz, K. Meehan, Plumbing Poverty: Mapping Hot Spots of Racial and Geographic Inequality in U.S. Household Water Insecurity, Ann. Am. Assoc. Geogr. 109 (2019) 1092–1109. https://doi.org/10.1080/24694452.2018.1530587.

[27] J. van Hoof, H.S.M. Kort, M.S.H. Duijnstee, P.G.S. Rutten, J.L.M. Hensen, The indoor environment and the integrated design of homes for older people with dementia, Build. Environ. 45 (2010) 1244–1261. https://doi.org/10.1016/j.buildenv.2009.11.008.

[28] E. Hood, Dwelling disparities: how poor housing leads to poor health., Environ. Health Perspect. 113 (2005) A310–7. https://doi.org/10.1289/ehp.113-a310.

[29] S.M. Joshi, The sick building syndrome., Indian J. Occup. Environ. Med. 12 (2008) 61–64. https://doi.org/10.4103/0019-5278.43262.

[30] E.G. Krug, L.L. Dahlberg, J.A. Mercy, A. Zwi, R. Lozano, The world report on violence and health, Lancet. 360 (2002) 1083–1088. https://doi.org/https://doi.org/10.1016/S0140-6736(02)11133-0.

[31] Centers for Disease Control and Prevention and Health Resources and Services Administration (CDC), The Social-Ecological Model: A framework for prevention, Violence Prev. (2022) 22–23. https://www.cdc.gov/violenceprevention/about/social-ecologicalmodel.html (accessed July 18, 2022).

[32] S. Ahmed, R. Muhammod, Z.H. Khan, S. Adilina, A. Sharma, S. Shatabda, A. Dehzangi, ACP-MHCNN: an accurate multi-headed deep-convolutional neural network to predict anticancer peptides., Sci. Rep. 11 (2021) 23676. https://doi.org/10.1038/s41598-021-02703-3.

[33] S. Hochreiter, J. Schmidhuber, Long Short-Term Memory, Neural Comput. 9 (1997) 1735–1780. https://doi.org/10.1162/neco.1997.9.8.1735.

[34] A. Deonarine, G. Lyons, C. Lakhani, W. De Brouwer, Identifying Communities at Risk for COVID-19–Related Burden Across 500 US Cities and Within New York City: Unsupervised Learning of the Coprevalence of Health Indicators, JMIR Public Heal. Surveill. 7 (2021). https://doi.org/10.2196/26604.

[35] A. Jamshidi, J.-P. Pelletier, J. Martel-Pelletier, Machine-learning-based patientspecific prediction models for knee osteoarthritis., Nat. Rev. Rheumatol. 15 (2019) 49–60. https://doi.org/10.1038/s41584-018-0130-5.

[36] Y. Lecun, Y. Bengio, G. Hinton, Deep learning, Nature. 521 (2015) 436–444. https://doi.org/10.1038/nature14539.

[37] J.A.M. Sidey-Gibbons, C.J. Sidey-Gibbons, Machine learning in medicine: a practical introduction, BMC Med. Res. Methodol. 19 (2019) 64. https://doi.org/10.1186/s12874-019-0681-4.

[38] M. Canizo, I. Triguero, A. Conde, E. Onieva, Multi-head CNN–RNN for multi-time series anomaly detection: An industrial case study, Neurocomputing. 363 (2019) 246–260. https://doi.org/https://doi.org/10.1016/j.neucom.2019.07.034.

[39] E.D. Gennatas, J.H. Friedman, L.H. Ungar, R. Pirracchio, E. Eaton, L.G. Reichmann, Y. Interian, J.M. Luna, C.B. Simone, A. Auerbach, E. Delgado, M.J. van der Laan, T.D. Solberg, G. Valdes, Expert-augmented machine learning, Proc. Natl. Acad. Sci. 117 (2020) 4571–4577. https://doi.org/10.1073/pnas.1906831117.

[40] T. Lesort, V. Lomonaco, A. Stoian, D. Maltoni, D. Filliat, N. Díaz-rodríguez, Continual learning for roboticslJ: Definition, framework, learning strategies, opportunities and challenges, 58 (2020) 52–68. https://doi.org/10.1016/j.inffus.2019.12.004.

[41] G.M. van de Ven, A.S. Tolias, Three scenarios for continual learning, ArXiv Prepr. ArXiv1904.07734. (2019). http://arxiv.org/abs/1904.07734.

[42] A. Lee, F.F.K. Cheng, D. Chow, 葵青健康城市及安全社區 –「社區診斷」調查計劃, 2004.

[43] J. Song, L. Zhang, G. Xue, Y. Ma, S. Gao, Q. Jiang, Predicting hourly heating load in a district heating system based on a hybrid CNN-LSTM model, Energy Build. 243 (2021) 110998. https://doi.org/10.1016/j.enbuild.2021.110998.

[44] A.P. Bradley, The use of the area under the ROC curve in the evaluation of machine learning algorithms, Pattern Recognit. 30 (1997) 1145–1159. https://doi.org/https://doi.org/10.1016/S0031-3203(96)00142-2.

[45] B. Yu, K. Kumbier, Veridical data science, Proc. Natl. Acad. Sci. U. S. A. 117 (2020) 3920–3929. https://doi.org/10.1073/pnas.1901326117.

[46] J. Merlo, B. Chaix, M. Yang, J. Lynch, L. Råstam, A brief conceptual tutorial of multilevel analysis in social epidemiology: Linking the statistical concept of clustering to the idea of contextual phenomenon, J. Epidemiol. Community Health. 59 (2005) 443–449. https://doi.org/10.1136/jech.2004.023473.

[47] H.M. Krumholz, R.G. Brindis, J.E. Brush, D.J. Cohen, A.J. Epstein, K. Furie, G. Howard, E.D. Peterson, S.S. Rathore, S.C. Smith, J.A. Spertus, Y. Wang, S.L.T. Normand, Standards for statistical models used for public reporting of health outcomes: An American Heart Association scientific statement from the Quality of Care and Outcomes Research Interdisciplinary Writing Group, Circulation. 113 (2006) 456–462. https://doi.org/10.1161/CIRCULATIONAHA.105.170769.

[48] M. Ding, Y. Wei, A conceptual framework for quantitatively understanding the impacts of floods/droughts and their management on the catchment’s social-ecological system (C-SES), Sci. Total Environ. 828 (2022) 154041. https://doi.org/https://doi.org/10.1016/j.scitotenv.2022.154041.

[49] Y.-B. Wen, Z.-R. Huang, Y.-F. Tang, D.-R. Li, Y.-J. Zhang, F.-Y. Zhao, Air exchange rate and pollutant dispersion inside compact urban street canyons with combined wind and thermal driven natural ventilations: Effects of non-uniform building heights and unstable thermal stratifications, Sci. Total Environ. 851 (2022) 158053. https://doi.org/https://doi.org/10.1016/j.scitotenv.2022.158053.

[50] D.P. Smith, O. Oechsle, M.J. Rawling, E. Savory, A.M.B. Lacoste, P.J. Richardson, Expert-Augmented Computational Drug Repurposing Identified Baricitinib as a Treatment for COVID-19., Front. Pharmacol. 12 (2021) 709856. https://doi.org/10.3389/fphar.2021.709856.

[51] A. Chou, A. Torres-Espin, N. Kyritsis, J.R. Huie, S. Khatry, J. Funk, J. Hay, A. Lofgreen, R. Shah, C. McCann, L.U. Pascual, E. Amorim, P.R. Weinstein, G.T. Manley, S.S. Dhall, J.Z. Pan, J.C. Bresnahan, M.S. Beattie, W.D. Whetstone, A.R. Ferguson, Expert-augmented automated machine learning optimizes hemodynamic predictors of spinal cord injury outcome., PLoS One. 17 (2022) e0265254. https://doi.org/10.1371/journal.pone.0265254.

[52] L. Moreno-Merino, H. Aguilera, A. de la Losa Román, Are bottled mineral waters and groundwater for human supply different?, Sci. Total Environ. 835 (2022) 155554. https://doi.org/https://doi.org/10.1016/j.scitotenv.2022.155554.

[53] S. Juneja, P. Shahabuddin, Chapter 11 Rare-Event Simulation Techniques: An Introduction and Recent Advances, Handbooks Oper. Res. Manag. Sci. 13 (2006) 291–350. https://doi.org/10.1016/S0927-0507(06)13011-X.

[54] C. Ma, X. Li, Z. Zhao, F. Liu, K. Zhang, A. Wu, X. Nie, Understanding the dynamics of pandemic models to support predictions of COVID-19 transmission: Parameter sensitivity analysis of the SIR-type model, IEEE J. Biomed. Heal. Informatics. 26 (2022) 2458–2468. https://doi.org/10.1109/JBHI.2022.3168825.

