## appendix for "Quantifying the Risk of General Health and Early COVID-19 Spread in Residential Buildings with Deep Learning and Expert-augmented Machine Learning"

Author Names and Affiliations

Jingjing GUAN^†,1,2^,

Eman LEUNG^†,*,1^,

Kin On KWOK^1,3,4,5^,

Chi Tim HUNG^1^,

Albert LEE^1,6^,

Ka Chun CHONG^1^,

Carrie Ho Kwan YAM^1^,

Clement KM. CHEUNG^7^,

Hendrik TIEBEN^8^,

Hector W.H. TSANG^6^,

Eng-kiong YEOH^1^

^†^Joint first authorship

*Corresponding author: Eman Leung Tel: +852 2252 8703, Fax: +852 2145 7489, JC School of Public Health and Primary Care, CUHK. Room 202, School of Public Health Building, Prince of Wales Hospital, Shatin, New Territories, Hong Kong

^1^JC School of Public Health and Primary Care, The Chinese University of Hong Kong, School of Public Health Building, Prince of Wales Hospital, Shatin, New Territories, Hong Kong Special Administrative Region, China.

^2^EPITELLIGENCE, Hong Kong Special Administrative Region, China.

^3^Stanley Ho Centre for Emerging Infectious Diseases, The Chinese University of Hong Kong, Postgraduate Education Centre, Prince of Wales Hospital, Shatin, New Territories, Hong Kong Special Administrative Region of China.

^4^Hong Kong Institute of Asia-Pacific Studies, Room 507, 5/F, Esther Lee Building, Chung Chi College, The Chinese University of Hong Kong, Shatin, New Territories, Hong Kong Special Administrative Region of China.

^5^Shenzhen Research Institute, The Chinese University of Hong Kong, 2001 Longxiang Boulevard, Longgang District, Shenzhen, China.

^6^Department of Rehabilitation Sciences, The Hong Kong Polytechnic University, Hung Hom, Hong Kong Special Administrative Region, China.

^7^The University of Hong Kong, Pokfulam, Hong Kong Special Administrative Region, China

^8^School of Architecture, Lee Shau Kee Architecture Building, The Chinese University of Hong Kong, Shatin, New Territories, Hong Kong Special Administrative Region, China.

**Appendices: Calculation of Feature Importance**

The deep learning model presented here is characterized by a multi-layered input, which includes 1) an embedding layer representing the number of the unique value of all features; 2) the concatenation of Long Short Term Memory (LSTM) layers, in which each LSTM layer represents the unique contribution of a feature where its values were mapped as perceptrons of the respective layer; and 3) in addition to individual LSTM layers, convolutional layers were constructed to reflect the interaction between layers to reflect the hierarchical relationship among features that represent different aspects of one’s social ecology.

Two sets of weights could be extracted for each feature from the embedding layer and the LSTM layer. The product of these two sets of weights formed a matrix of a few vectors. Each vector corresponded to a unique value of the inputted feature. To evaluate the contribution of each actual value of a feature, we compared its corresponding vector to two vectors (0s and 1s) of the same length.

Without loss of generality, we denoted the vector of an actual value as *v*.

The first vector was a zero vector *z* whose elements were all zeros. The zero vector *z* represented an actual value that was not selected by the deep learning model (“forgotten” from the LSTM), and hence the actual value had no effects on the outcome variable. The Euclidean distance between the *v* and *z* was computed. The greater the Euclidean distance, the greater the effect of the actual value on the outcome variable.

The second vector was a unit vector *u* whose elements equaled 1. We computed the Cosine distance between *v* and *u*. The unit vector *u* represented that an actual value had a completely positive collinear association with the outcome variable. The Cosine distance measured the angle between the vector *v*  of the actual value of a feature and the unit vector *u*. Such an angle was actually the angle between the effect of the actual value and the outcome variable. Ranging between -1 and 1, a negative Cosine distance means a negative association between the actual value and the outcome variable, while a positive Cosine distance means a positive association. When the Cosine distance equals -1 or 1, the actual value has a complete collinear association with the outcome variable.

We further derived the feature importance as the collinearity between the vector *v* and the outcome variable, which was equivalent to the product of the Euclidean distance and Cosine distance. Feature importance could be interpreted in the same way as a regression coefficient, where a positive value corresponds to a positive effect, a negative value corresponds to a negative effect, and a greater magnitude means a greater effect size.

Graphically, the zero vector represents the origin of a hyperspace, as shown in the figure below.

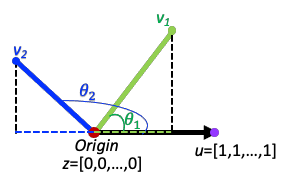

The unit vector is the purple point located at the horizontal direction of the outcome variable represented by the black arrow.

The feature importance of vector *v*_1_ is its projection on the horizontal axis, represented by the green dashed line.

The feature importance of vector *v*_2_ is also its projection on the horizontal axis, represented by the blue dashed line. The Euclidean distance between the zero vector respectively with the vectors *v*_1_ and *v*_2_  is represented by the green solid line and blue solid line, while the Cosine distance between the unit vector respectively with vectors v_1_ and v_2_ is represented by the cosine of $\theta_{1}$ and $\theta_{2}$.
